# Scaling-up tuberculosis preventive therapy to over five-years old HIV-negative contacts of bacteriologically confirmed TB cases – lessons from a tuberculosis preventive therapy surge activity in Zambia

**DOI:** 10.1101/2025.11.26.25341134

**Authors:** Phallon Blessing Mwaba, Innocent Mwaba, Minyoi Mubita Maimbolwa, Chitalu Chanda, Muhammad Mputu, Chola Lumbwe, Kevin Zimba, Nancy Kasese Chanda, Rhehab Chimzizi, Angel Mubanga, Monde Muyoyeta, Mary Kagujje

## Abstract

**Background:** While tuberculosis preventive therapy (TPT) coverage among people living with HIV in Zambia has improved, uptake among HIV-negative contacts aged ≥5 years remains low. Despite some evidence that TPT can reduce TB incidence by up to 60% in HIV-negative individuals, there is a paucity of evidence on programmatic rollout to this population in Zambia and the region. We present programme data from a TPT surge aimed at increasing uptake among ≥5 years old HIV-negative household TB contacts.

**Methods:** The National TB Programme, with support from the TB Local Organizations Network, conducted a TPT surge in 89 high TB-burden facilities across eight project supported provinces in Zambia from October 2024 to 15^th^ January 2025. After a thorough planning phase, multidisciplinary teams conducted field visits for contact tracing and initiated TPT for eligible household contacts of bacteriologically confirmed TB, regardless of HIV status or age. Facility-based TPT initiation was provided for walk-in clients. We collected aggregate data from facility registers and analyzed it descriptively.

**Results:** A total of 21,890 HIV-negative contacts were screened: 21,482 (98.1%) eligible for TPT. Among these, 15,711(74.6%) were initiated on TPT. Of those screened, 19,607 (91.8%) were >5 years old; 19,345(98.7%) of them were eligible, and 14,000 (72.4%) were initiated on TPT. Overall, 61.2% (n=9432) were initiated on three months Isoniazid + Rifapentine, followed by six months Isoniazid(32.6%, n=5125); one month Isoniazid and Rifapentine (4.1%, n=638) and on three months Isoniazid and Rifampicin (1.7%, n=272).

**Conclusion:** The Surge demonstrated that high TPT coverage among ≥5 years old HIV-negative TB contacts is achievable with targeted implementation, stakeholder engagement, and community-driven approaches. These findings underscore the importance of expanding routine tuberculosis preventive therapy delivery to include all high-risk contacts, regardless of HIV status or age, to accelerate TB prevention efforts in high-burden settings like Zambia.

## Introduction

Zambia is among the top 30 high TB burden countries globally, with a tuberculosis (TB) incidence of 283/100,000 population (1). The country has made remarkable progress in lowering the TB burden in people living with HIV (PLHIV) with the TB incidence reducing from 205 per 100,000 in 2018 to 91 per 100,000 in 2023 in this population (2). However, there has been a shift of the TB burden to the HIV negative population. Nearly 70% of the total TB notifications in Zambia in 2023 were HIV negative (1).

In 2018, Zambia’s Ministry of Health (MoH) adopted the World Health Organisation (WHO) recommendation to extend TB preventive Therapy (TPT) delivery beyond PLHIV and under 5 contacts of TB patients to include HIV negative contacts aged older than 5 years of bacteriologically confirmed TB patients and other high risk groups for TB (3). Similar to other settings, and in spite of strong evidence that TPT can reduce TB incidence by up to 60% in HIV-negative individuals (4–6), its implementation has been slow and variable (7). Between 2018 and 2021, Zambia made significant progress in TPT implementation, achieving a 90% TPT uptake among PLHIV (8). However, the coverage among HIV negative household contacts to bacteriologically confirmed TB patients remains much lower, at 55% (1), and these mostly comprise children below 5 years of age.

To address the current gaps in TPT implementation, the national TB program developed the implementation roadmap for TPT, aimed at saturating and sustaining TPT in PLHIV, accelerating TPT uptake in under 5 contacts to TPT cases and increasing focus on other high-risk categories such as all contacts to bacteriologically confirmed TB patients regardless of age and HIV status (9). In alignment with the roadmap, The United States Government funded Tuberculosis Local Organisations Network (TBLON) project implemented a TPT Surge to scale up TPT to HIV negative contacts to bacteriologically confirmed TB patients aged >5 years. We present programme data from this surge to demonstrate the feasibility and acceptability of TPT among HIV-negative household contacts aged ≥5 years, with the aim of informing programmatic scale-up.

## Methodology

### Intervention design and setting

We implemented a TPT surge in routine settings between 1^st^ October 2024 and 15^th^ January 2025 in 89 high TB burden public health facilities in eight out of Zambia’s ten provinces (Southern, Lusaka, Central, Muchinga, Copperbelt, Northern, Luapula and Northwestern). These facilities receive support from TBLON. In each province, we selected sites contributing 80% to provincial TB notifications: a mix of primary, secondary and tertiary level health facilities. Routinely, household contacts of bacteriologically confirmed TB are symptomatically screened and investigated for TB. Those diagnosed with TB are put on treatment while mainly eligible PLHIV contacts (not currently or recently given TPT) and <5 children established to have no active TB disease are initiated on TPT.

### Surge Intervention

This intervention was guided by the Exploration, Preparation, Implementation, and Sustainment (EPIS) framework (10,11), which structures intervention planning across sequential phases and contextual layers. In the exploration stage, we used a needs assessment to identify gaps and contextual factors affecting uptake. In the preparation stage, we developed strategies to leverage facilitators and address barriers, ensuring readiness for implementation. During the implementation stage, we delivered activities with attention to fidelity and adaptability. For the sustainment stage, we supported integration of effective interventions into routine practice. In this paper, we focus on the pre-surge (exploration and preparation) and intra-surge (implementation) phases of the surge, highlighting their role in guiding planning and operationalization within the target context.

#### Pre-Surge Activities

We developed a TPT surge concept note in consultation with the National Tuberculosis and Leprosy Program (NTLP) and set targets. The targets were set a priori with the assumption that refusal rate would be higher among HIV negative contacts aged >5 years. We targeted TPT initiation for 85% of eligible <5 years old contacts and at least 2,000 HIV negative household contacts to bacteriologically confirmed TB patients aged ≥5 years.

We conducted community engagement activities including media campaigns, door-to-door sensitization, and community meetings. We engaged MoH staff, community-based volunteers, and local gatekeepers to promote TPT uptake and address myths and misconceptions in the community. We also assessed and mobilized TPT commodities to ensure a steady supply across facilities during the surge.

We also oriented health care workers and community-based volunteers (CBVs) in TPT guidelines and surge targets to ensure a uniform approach. Additionally, we distributed copies of TPT guidelines and job aids to all participating sites as reference documents. To facilitate surge monitoring, we generated and piloted a DHIS2 tracker-based digital reporting tool.

#### Intra-Surge Activities

Two groups of contacts of bacteriologically confirmed index TB cases were targeted: 1) contacts of TB patients diagnosed within three months prior to the surge 2) contacts of newly diagnosed TB patients. The surge primarily utilized a community-based approach, with multidisciplinary teams (each team comprising a nurse/clinician, a Community Health Worker and a Community Mobilization Officer where necessary), coordinated by local health facility staff, conducting home visits for contact investigation and community TPT initiation. We also provided TPT to walk-in clients at health facilities. We used symptom screening to rule out TB as recommended for high TB burden areas (5). We sustained demand generation activities during the surge through radio programmes and community engagement meetings addressing emerging reasons for TPT refusals.

We conducted daily monitoring and supervision to ensure surge implementation with fidelity and tracking of key indicators (TPT eligibility, initiation rates, refusal rates, and commodity stock levels). Furthermore, we held weekly progress review meetings and shared reports with key stakeholders: the MoH and USAID. We documented all activities, using a journal, to track implementation components potentially key to effective TPT rollout to the HIV negative at-risk populations aged 5 years and above in routine practice.

### Data collection methods

Data collection spanned the TPT surge implementation period from 1^st^ October 2024 to 15^th^ January 2025. Facility based MoH staff routinely documented surge data in facility-based registers while project data associates collected and entered the aggregate data into the electronic DHIS2 based tool weekly. Additionally, we extracted routinely collected programme data from the Facility Information Management System (FIMS) from January to September 2024 to inform analysis of TPT initiation trends.

### Data Management and analysis

We extracted aggregate TPT data from the electronic data collection tool into excel and conducted descriptive analysis. We presented the annual aggregate data graphically to show the trends of TPT coverage among contacts to bacteriologically confirmed TB contacts between January and December 2024. The team analyzed intra-surge TPT initiation including TPT initiation stratified by age category, sex, HIV status and TPT regimen and presented it in the form of counts and proportions. Furthermore, information from decline forms was extracted to understand the common reasons for refusal of TPT.

### Ethical Considerations

Considering this was based on routinely collected programme data, there was no additional ethics approval obtained beyond the pre-approval obtained from University of Zambia Biomedical Research and Ethics Committee (UNZA BREC), Lusaka, Zambia (Reference number 1773–2021) for the use of routinely collected programme data.

## Results

Overall, 22,158 household contacts were enlisted during the surge, of whom 21,890 (98.8%) completed pre-TPT screening (Fig 1). Among those screened, 21,482 (98.1%) were found eligible for TPT. Of the eligible population, 449 (2.1%) declined TPT, while 5,322 (24.8%) accepted TPT but had not initiated treatment by the end of the surge due to unavailability of their preferred regimen. The remaining 15,711 (73.1%) contacts were initiated on TPT, the majority (98.0%; n=15,401) of whom were HIV-negative.

**Fig. 1.**
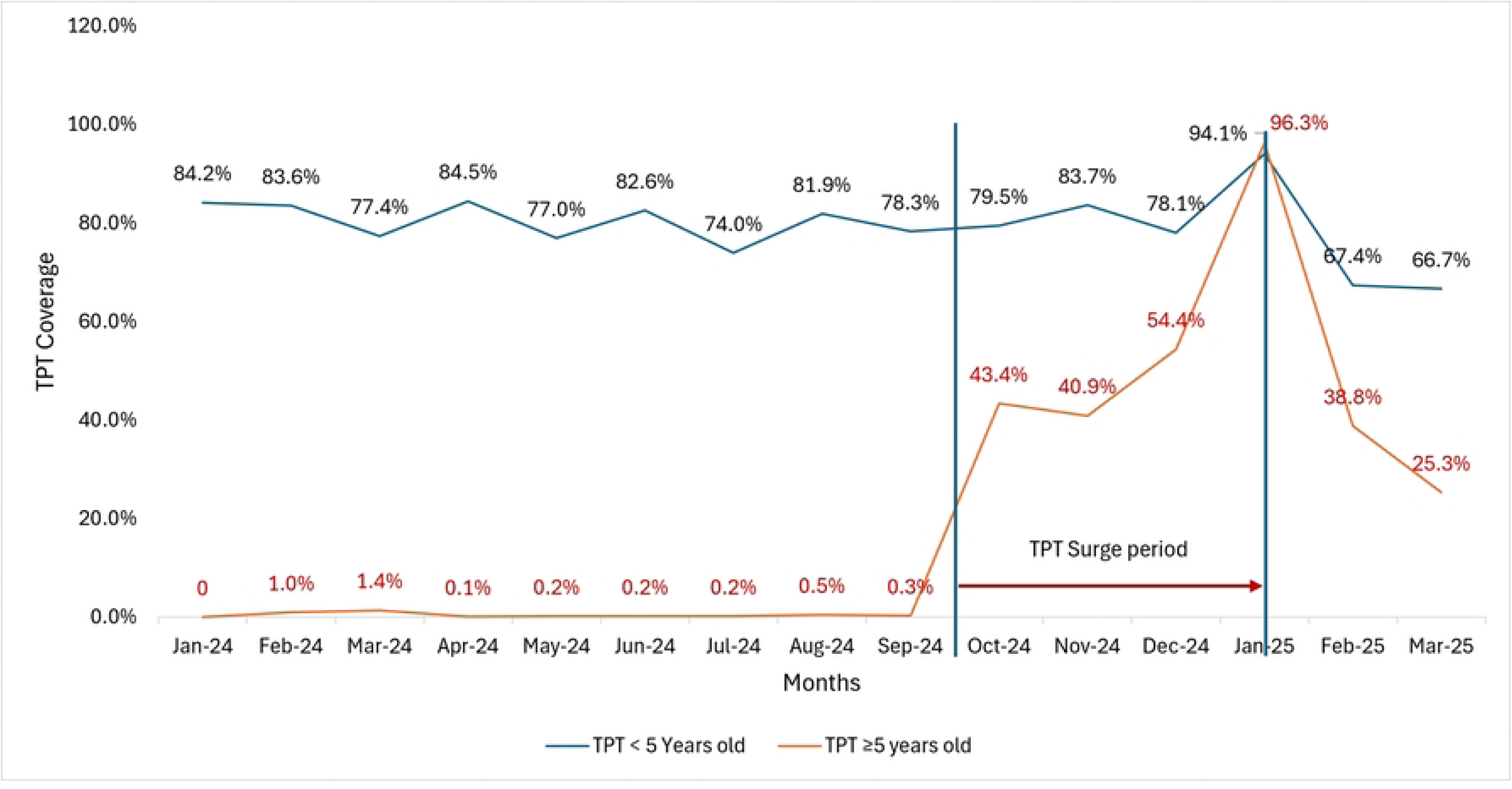
TPT surge flow chart for TB contacts.

Of the 15,401 HIV negative contacts initiated on TPT, 1,401 (9.1%) were aged <5 years, while 14,000 (90.9%) were ≥ 5 years old (Table 1). Females accounted for 52.7% of those initiated on TPT. Sites of TPT initiation included rural health centres (9.1%, n=1405), urban health centres (33.2%, n=5111), level one hospitals (39.1%, n=6026), second level hospitals (13.2%, n=2038) and tertiary level hospitals (5.3%, n=821).

**Table 1.** Characteristics of HIV negative Contacts initiated on TPT (N=15401)

| Variable | Number of Contacts n(%) |
| --- | --- |
| **Age group (years)** |  |
| 0-4 | 1401 (9.1) |
| 5-14 | 3414 (22.2) |
| 15-64 | 10139 (65.8) |
| $\geq 65$ | 447 (2.9) |
| **Sex, n (%)** |  |
| Male | 7280 (47.3) |
| Female | 8121 (52.7) |
| **TPT initiating facility type, n (%)** |  |
| Third Level | 821(5.3) |
| Second Level Hospital | 2038(13.2) |
| First Level Hospital | 6026(39.1) |
| Urban HC | 5111(33.2) |
| RHC | 1405(9.1) |

Among HIV negative household contacts elicited (n = 21,353), we screened 21,091 (99.0%) for TB (S1 Table). Of those screened, we identified 1,452 (6.9%) presumptive TB cases among whom 39 (2.7%) were diagnosed with TB and initiated on treatment. The rest of the contracts were eligible for TPT out of whom 15401(73.2%) were initiated on TPT. Specifically for the 19,345 over 5 years old HIV negative contacts eligible for TPT, 72.3% (n =14, 000) were provided with TPT.

**Table 2:** Age aggregated TPT cascade for HIV negative contacts to Bacteriologically confirmed TB (n (%)

Overall, most contacts (n=9676, 62.8%) were initiated on three months Isoniazid + Rifapentine (3HP), followed by six months Isoniazid (6H) (n=5125, 32.9%); one month Isoniazid and Rifapentine (1 HP) (n=638, 4.1%) and the least on three months Isoniazid and Rifampicin (3HR) (n=272(1.7%) (Table 3).

**Table 3.**
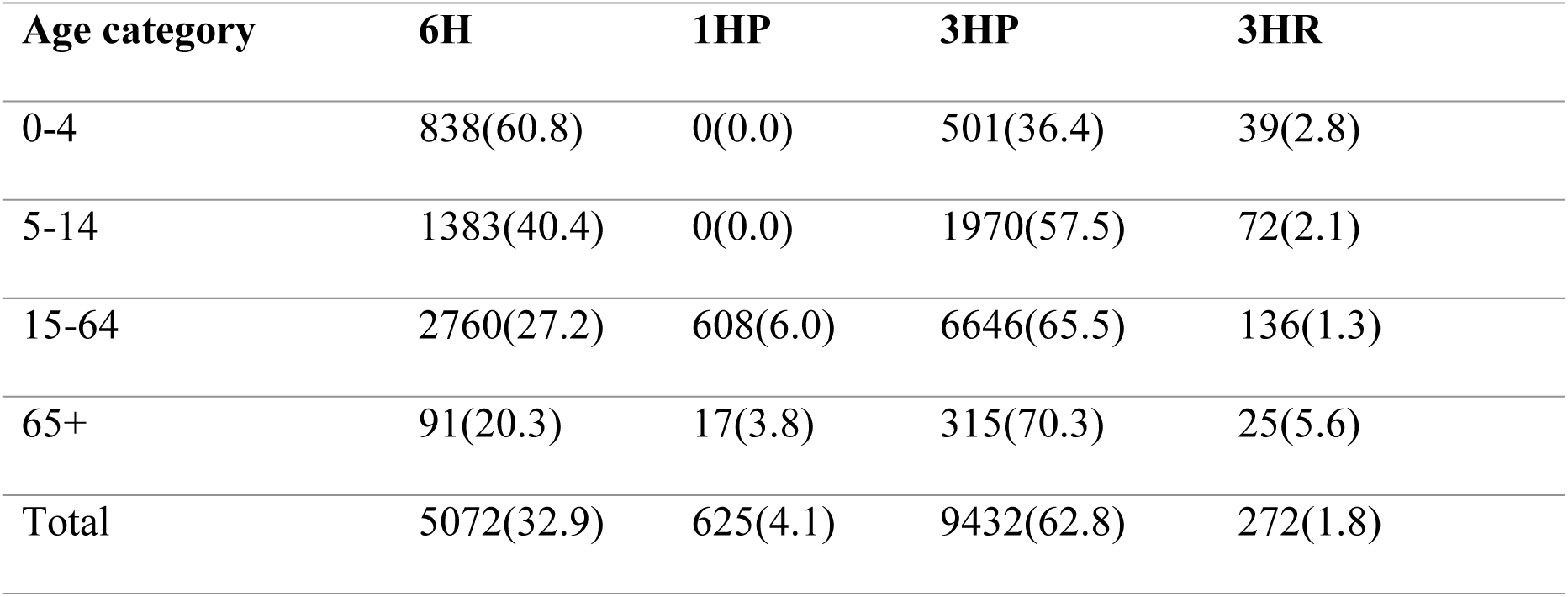
Types of TPT regimens initiated for eligible TB contacts, disaggregated by age, n(%)

| Age category | 6H | 1HP | 3HP | 3HR |
| --- | --- | --- | --- | --- |
| 0-4 | 838(60.8) | 0(0.0) | 501(36.4) | 39(2.8) |
| 5-14 | 1383(40.4) | 0(0.0) | 1970(57.5) | 72(2.1) |
| 15-64 | 2760(27.2) | 608(6.0) | 6646(65.5) | 136(1.3) |
| 65+ | 91(20.3) | 17(3.8) | 315(70.3) | 25(5.6) |
| Total | 5072(32.9) | 625(4.1) | 9432(62.8) | 272(1.8) |

Compared with pre-surge programme data, TPT coverage for children < 5 years old was high both pre- and intra-surge, ranging from 74.0% in July 2024 to a peak of 94.1% observed in January 2025 with minor fluctuations (Fig 2). Conversely, TPT coverage among HIV negative contacts aged ≥5 years ranged from 0% to 1.4% between January and September 2024. During the surge period (October 2024 to January 2025), TPT coverage among HIV-negative contacts increased sharply from a baseline of 0.3% (15/5,366) in September to 43.4% in October, 40.9% in November, 54.4% in December and ultimately peaking at 96.3% in January 2025.

**Fig. 2.**
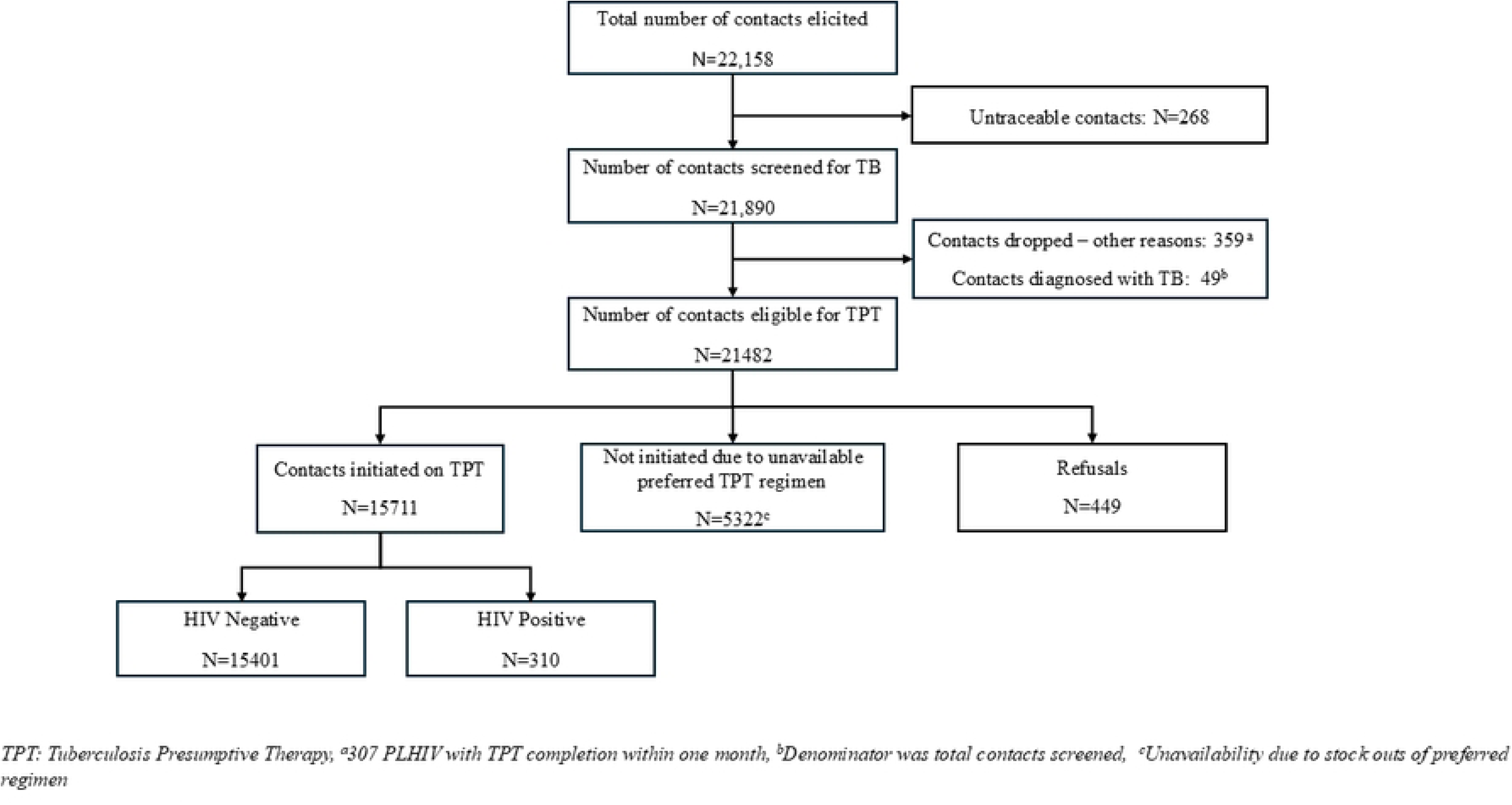
Pre- and intra-surge TPT Coverage trends in TB LON supported sites in 2024.

After the surge, TPT coverage among contacts aged ≥5 years declined to 38.8% in February and 25.3% in March. A similar downward trend was observed among contacts aged <5 years, whose coverage declined to 67.4% and 66.7% in February and March, respectively.

## Discussion

This analysis revealed that by the end of the surge implementation, TPT initiation among HIV-negative household contacts aged ≥5 years had increased from a baseline of 0.3% before the surge to 96.3%, exceeding historical national average and surpassing the WHO-recommended 85% coverage threshold among contacts (12,13). After the surge, a decline in TPT coverage was observed; mostly attributable to the impact of the United States Government stop work order: sudden withdrawal of support for TB activities. This is the first implementation study in Zambia and regionally to demonstrate feasibility and effectiveness of a surge approach in scaling up TPT coverage among HIV-negative household contacts to bacteriologically confirmed TB aged ≥5 years.

This intervention demonstrated that a well-planned, well-designed and executed TPT surge targeting all TB household contacts can improve uptake among HIV negative contacts aged ≥5 years: a population historically under-prioritized in TB prevention programming. The success of the TPT surge can be attributed to adequate planning, healthcare workers capacity building, close performance monitoring as well as community engagement and demand generation pre- and intra-surge. Activities such as health education through door-to-door sensitization, community meetings, and radio programmes potentially contributed to increased uptake by addressing low risk perceptions, fear of side effects, beliefs, myths and misconceptions and other documented barriers to TPT acceptance, especially among adults (14). Furthermore, integrating trained community-based volunteers and local champions into community engagement and service delivery may have improved trust and supported TPT acceptance. This aligns with growing evidence that community-based delivery models coupled with systematic contact tracing and decentralized service delivery, can improve access and uptake of TB services in high-burden settings (15–17) especially if delivered by adequately capacity built multi-disciplinary teams, as was done in this surge, and adequately resourced (4).

While global and national guidelines have long recommended provision of TPT to all contacts of bacteriologically confirmed TB regardless of HIV status (1,7,18), the focus in Zambia, like other low income countries, has remained skewed toward PLHIV and ≤5 years old children (19). Consequently, HIV-negative contacts ≥5 years old, despite constituting the majority of household contacts, have remained largely underserved (7). As a result, the TB burden among HIV-negative individuals has largely remained static or has declined at a much slower rate over the past decades compared to the PLHIV (2). Despite the overall TB burden remaining high among PLHIV (20), there is a decline in TB incidence in this group; a trend mainly attributable to the combined effect of antiretroviral therapy (ART) and TPT (1,21). Considering the documented effectiveness of TPT in reducing incident TB (4,5), improving coverage can help reduce TB morbidity and community spread among this population; critical ingredients for ending TB (4,7,22). Despite a lower risk of tuberculosis among HIV-negative (immunocompetent) individuals compared to those with HIV, our findings and recent literature suggest that the cost of failing to protect this group is substantial and under-appreciated. While PLHIV are not spared, strong immune responses during active disease in immunocompetent HIV negative people frequently result in structural lung damage (cavitation, fibrosis, and bronchiectasis) which may persist after cure and drive post-TB lung disease (PTLD) with about 20% to 45% of TB survivors exhibiting PTLD signs and symptoms among this population (23); significantly higher than among PLHIV (24,25).

Our findings challenge the assumption that low TPT uptake in this group is due to poor acceptability (26); instead suggesting that previous low coverage may be a function of system-level weaknesses such as ineffective TPT delivery approaches and a lack of prioritization rather than low acceptability among targets. This aligns with previous qualitative studies among health care workers in South Africa that established that HCWs’ negative attitudes and system weaknesses such as poor contact investigation strategies and resource shortages are key drivers of low TPT uptake among TB contacts (14). Therefore, more investment, system strengthening and resource investment are required to improve TPT uptake among all TB contacts regardless of age and HIV status.

The high initiation rate achieved during the surge (73.5%) demonstrates that substantially high acceptance rates are attainable when implementation barriers such as low-risk perception (14) are effectively addressed. This suggests that pre-surge concerns about potential poor uptake were overstated. Notably, the refusal rate observed among HIV-negative household contacts aged ≥5 years was only 2.1%, highlighting the potential acceptability of TPT in this demographic when interventions are well planned, executed with fidelity, community-focused and adequately resourced. This contrasts with earlier studies that reported markedly lower acceptability among HIV-negative household contacts aged ≥5 years, in some cases as low as 3% (1,27), where fear of side effects, misconceptions, and limited risk perception were cited as key barriers (14).

Although commodity shortages during the surge may have influenced regimen availability, the distribution of TPT regimens revealed a pronounced preference for the short-course rifapentine-based 3HP regimen (62.8 %), particularly among adults. This preference is supported by WHO’s 2020 recommendations, which grant 3HP strong endorsement due to its non-inferior efficacy, convenience of administration, and lower hepatotoxicity compared to longer isoniazid regimens (3, 20). Moreover, nearly 25% of the TB contacts could not commence TPT despite being willing to take it due to unavailability of the preferred shorter regimen. Studies have shown higher completion and adherence rates for 3HP compared to longer regimens like 6H (29,30). Recent systematic reviews of completion rates of 3HP relative to other treatment regimens for latent tuberculosis infection revealed that shorter, 3 to 4 months long, regimens were more likely to be completed than longer regimens (31,32). Real-world implementation in Uganda among PLHIV confirmed these findings: acceptance and completion exceeded 90 % across different delivery strategies (33). Collectively, these findings suggest that when supply permits, patients and providers strongly favor the shorter, more patient-friendly 3HP regimen. Service providers must therefore ensure reliable procurement and supply-chain resilience to maintain access to rifapentine-based TPT.

Even though Zambia’s national TB incidence remains high overall (283/100,000) (1), the notable recent decline in TB among PLHIV mainly attributed to ART and TPT scale-up in this population (1,21,34) can be replicated in the HIV negative population. Scaling up TPT nationally among HIV negative contacts to bacteriologically confirmed TB using the strategies tested in this surge could help flatten the TB incidence curve among this population by reducing community transmission, as previously modelled in other high-burden settings (5,21,35). Moreover, this initiative also presents a replicable model for other high-burden countries aiming to operationalize the WHO’s person-centered and community-oriented TB prevention guidance (3). The success of the Zambia TPT Surge in improving TPT coverage among HIV negative contacts aligns with global TB elimination goals outlined in the UN High-Level Meeting Declaration (36), which calls for inclusive, equitable, and context-appropriate delivery of preventive services to all at-risk populations, including HIV-negative individuals.

Despite the successes of this study, it did not lack limitation. This analysis relied on routinely collected aggregate data, which may be subject to reporting inaccuracies or underreporting and limit the analyses that could be conducted. Nonetheless, the stark increase in uptake following implementation provides strong suggestive before and after evidence of the intervention’s effectiveness and a proof of concept for scaling up TPT to HIV negative contacts >5 years old in Zambia and elsewhere.

## Conclusion

The TPT Surge demonstrated that with intentional design, staff capacity building, strong community engagement, and real-time data monitoring, it is feasible to achieve high TPT coverage among HIV-negative contacts aged ≥5 years in Zambia and beyond. These results support national and global calls for inclusive, person-centered TB prevention strategies that do not exclude older HIV-negative individuals. As Zambia moves to institutionalize its TPT roadmap, integrating surge elements into routine programming could accelerate progress toward TB elimination.

## Data Availability

All relevant data are within the paper and its Supporting Information files. Additional datasets generated and/or analyzed during the current study are available from the corresponding author upon reasonable request.

## Availability of Data and materials

All data will be submitted as part of this manuscript

## Authors Contributions

Data extraction was done by Innocent Mwaba

Data cleaning and analysis was done by Innocent Mwaba and Phallon Mwaba

The original draft of the manuscript was by Phallon Mwaba

Critical review of the manuscript was done by Chitalu Chanda, Muhammed Mputu, Minyoi Maimbolwa, Chola Lumbwe, Angel Mubanga, Nancy Chanda Kasese, Kevin Zimba, Mary Kagujje, and Monde Muyoyeta.

Supervision and validation were done by Monde Muyoyeta and Mary Kagujje

All authors reviewed the manuscript and agreed to publish.

## Acknowledgements

The authors would like to thank the staff at MoH health facilities who were involved in documenting data in registers from where data was collected.

